# # Treatment- and tissue-context-specific isotope labeling patterns in patient-derived lung cancer tissues: an independent reanalysis of public metabolomics data

**DOI:** 10.64898/2026.11.15.26368161

**Authors:** Zhijie Shang, Hong Geng

**Affiliations:** Department of Thoracic Surgery, Shanxi Bethune Hospital, Third Hospital of Shanxi Medical University, Shanxi Academy of Medical Sciences, Tongji Shanxi Hospital, Taiyuan 030032, Shanxi, China; Department of Thoracic Surgery, Yuci District People’s Hospital, Jinzhong 030600, Shanxi, China

**Author notes:** **Corresponding author:** Zhijie Shang, Department of Thoracic Surgery, Shanxi Bethune Hospital, 99 Longcheng Street, Taiyuan 030032, Shanxi, China.

**Keywords:** lung cancer, isotope-resolved metabolomics, pembrolizumab, β-glucan, pentose phosphate pathway, nucleotide metabolism, reproducibility, secondary analysis

## Abstract

Publicly deposited stable-isotope-resolved metabolomics data can be reanalyzed independently, but small patient-derived experiments may produce treatment effects with limited statistical resolution. We reanalyzed a public isotope-resolved metabolomics dataset from patient-derived lung cancer tissue cultures to quantify treatment- and tissue-specific labeling patterns and their uncertainty.

We used 38 tissue samples from two patients: matched tumor and non-tumor slices from UK131 treated with vehicle or pembrolizumab and traced with U-¹³C₆-glucose or U-¹³C₅,¹⁵N₂-glutamine (n = 3 per cell), and a pembrolizumab × WGP β-glucan factorial experiment in UK2035 tumor tissue (n = 3/3/3/2). We recomputed protein-normalized pools, isotopologue distributions, atom-percent excess and pathway summaries. Effects were estimated with log₂-scale linear models, Hedges’ g, and exact randomization tests that enumerated all assignments compatible with each design; Benjamini–Hochberg q values were calculated within endpoint families. Sixteen macrophage samples were examined only descriptively.

In UK131 tumor tissue, labeled non-oxidative pentose phosphate pathway intermediates increased 1.84-fold (model P = 0.036; exact tumor P = 0.10; exact interaction P = 0.16; q = 0.32). Weighted isotopic scrambling increased 1.31-fold (exact interaction P = 0.04; q = 0.20), and glutamine-derived glutathione diversion increased 1.66-fold (exact interaction P = 0.09; q = 0.22). In UK2035, the combination increased the labeled IMP/(ATP+GTP+UTP) ratio 3.64-fold (model interaction P = 3.2 × 10⁻⁴; exact P = 0.016; q = 0.097). No endpoint survived Benjamini–Hochberg correction at q < 0.05 in the exact analyses.

The reanalysis reproduced the direction of the source study’s principal labeling patterns but showed that their statistical support is weaker than model-based P values imply. These results should be treated as hypothesis-generating estimates for larger patient-derived studies, not as evidence of a validated metabolic bottleneck.

## Introduction

Immune checkpoint blockade has changed the treatment landscape for non-small cell lung cancer, but response remains heterogeneous [1–3]. Tumor and immune cells compete for glucose, glutamine and nucleotide precursors, and metabolic adaptation may influence both antitumor immunity and resistance to immunotherapy [3–7]. Stable-isotope-resolved metabolomics (SIRM) can distinguish changes in metabolite abundance from changes in the fate of imported carbon and nitrogen [8–13]. In particular, U-¹³C₆-glucose tracing reports oxidative and non-oxidative pentose phosphate pathway (PPP) labeling as well as glycogen-related sugar phosphate cycling, whereas U-¹³C₅,¹⁵N₂-glutamine tracing reports entry into the tricarboxylic acid (TCA) cycle, aspartate synthesis and glutathione (GSH) metabolism [11–17].

The source study of the dataset analyzed here used patient-derived organotypic tissue cultures to show that pembrolizumab activated innate immune programs in lung cancer tissue, altered glucose and glutamine handling in one patient, and that pembrolizumab plus the β-glucan whole glucan particle (WGP) suppressed central metabolism in a second patient [8,9]. β-glucan can also reprogram myeloid cells through trained-immunity and immunometabolic programs that differ from checkpoint-receptor blockade [18,19,20,21]. Those experiments were explicitly patient-specific: they involved two tumor patients, small numbers of biological replicates, a single labeling time point and no independent validation cohort. The source report therefore provided valuable proof-of-principle evidence but did not establish whether its quantitative metabolic patterns were reproducible under an independent analysis or how much statistical uncertainty surrounded the reported effects.

Independent reanalysis of public SIRM data is useful for three reasons. First, it tests whether reported patterns can be recovered from deposited raw tables when pools, isotopologue distributions and enrichment measures are recomputed. Second, it makes analysis assumptions visible, including normalization, endpoint construction and multiple-testing decisions. Third, it permits exact randomization inference, which is particularly relevant when treatment cells contain only two or three samples and conventional parametric tests may be anti-conservative [22–25]. Such a reanalysis cannot create new biological replicates or establish flux, but it can distinguish a reproducible directional signal from an overconfident summary of a fragile experiment.

We therefore performed an independent, data-level reanalysis of the public deposit associated with the source study [8,9]. Our objectives were to (i) quantify pembrolizumab-associated glucose and glutamine labeling patterns in matched tumor and non-tumor tissue from UK131; (ii) quantify the pembrolizumab × WGP factorial structure in UK2035 tumor tissue; (iii) compare conventional model-based inference with exact randomization tests; and (iv) define the boundary between hypothesis-generating metabolic patterns and validated mechanisms. We did not attempt to replace the source study’s immune profiling or to infer clinical response.

## Materials and methods

### Data source and experimental design

We used the Dryad deposit doi:10.5061/dryad.n5tb2rbwc, associated with the published study by Fan et al. [8,9]. The deposit contains five Microsoft Excel workbooks, a README file and the source article. The reanalysis was performed in September 2026 from the deposited tables without access to unpublished data.

The primary analysis used 38 tissue samples from two patients. For UK131, organotypic tissue slice cultures from tumor (CA) and non-tumor (NC) lung tissue were treated with vehicle or pembrolizumab (40 μg/mL as encoded in the sample identifiers) for 24 hours in the presence of either U-¹³C₆-glucose or U-¹³C₅,¹⁵N₂-glutamine, with n = 3 per tissue × treatment cell (24 samples). Eight glucose-tracing metabolites and 10 glutamine-tracing metabolites passed the quality-control criteria used here.

For UK2035, metastatic lung cancer tissue was analyzed in a 2 × 2 factorial design. Block CA contained three vehicle-treated samples only. Block CA2 contained vehicle (n = 3), pembrolizumab (n = 3), WGP β-glucan (n = 3) and pembrolizumab + WGP (n = 2), all traced with U-¹³C₆-glucose (11 samples). Because the two blocks differed systematically in absolute pool scale, all factorial inference was restricted to CA2, which contained all four treatment groups. Twenty-four metabolites were quantified in this experiment.

The deposit also contains 16 monocyte-derived macrophage samples from donor UK096 (M0, M1, M2 and M2 + WGP; n = 2 per condition). Because these samples had no tumor tissue, no pembrolizumab treatment and only two replicates per condition, they were used exclusively for the exploratory cross-system comparison reported in the Supporting Information. They did not contribute to any primary estimate or conclusion.

### Data extraction and quality control

The deposited workbooks were parsed at the level of sample × metabolite × isotopologue. Blank rows, unlabeled standards, bulk runs and embedded header rows were removed. Exact duplicate rows and duplicated sample–metabolite–isotopologue entries were removed. The final design-filtered dataset contained 6,366 quantified sample–metabolite–isotopologue rows.

The primary quantity was the deposited protein-normalized metabolite amount (Amount_ProteinAdj_uMol_g_protein_SequenceBased), expressed as μmol metabolite per g protein. The alternative ratio-based normalization was retained for a calibration-sensitivity check. NADPH was excluded because no authentic standard was available, and quinolinic acid was excluded because it was quantified against a standard in only part of the UK131 glutamine panel and produced non-physiological concentrations.

Replicate variability was summarized as the within-cell coefficient of variation (CV) for metabolites with at least two samples in a treatment group. Coverage was summarized separately for each experiment because the metabolite panels were experiment-specific rather than missing at random. One UK131 tumor sample in the glutamine experiment covered only part of the panel; no values were imputed.

### Isotope and pathway measures

For each sample × metabolite, total pool was calculated as the sum of isotopologue amounts. The labeled amount was total pool minus the M+0 amount. Atom-percent excess (APE) for carbon was calculated as Σ(i × amountᵢ)/(nC × pool), where i is the number of labeled carbon atoms and nC is the carbon number of the metabolite. Nitrogen-percent excess was calculated analogously. For glucose experiments, isotopic scrambling was defined as labeled amount minus the fully labeled isotopologue amount, and the scrambling fraction was scrambling divided by labeled amount.

Pathway scores were sums of labeled amounts across metabolites assigned to predefined pathways. They are labeling summaries, not flux estimates [13]. The following composite indices were calculated as arithmetic summaries of the reanalysis:

- Non-oxidative PPP share: labeled non-oxidative PPP metabolites divided by the sum of labeled glucose-6-phosphate, 6-phosphogluconate, non-oxidative PPP metabolites and glycogen-branch metabolites.
- Oxidative PPP share: labeled 6-phosphogluconate divided by the same denominator.
- G6PD partition: labeled 6-phosphogluconate divided by labeled glucose-6-phosphate plus labeled 6-phosphogluconate.
- Weighted isotopic scrambling: labeled-amount-weighted mean scrambling fraction across non-oxidative PPP metabolites.
- GSH diversion: labeled GSH divided by labeled glutamate plus labeled GSH.
- Nucleotide bottleneck index (NBI): labeled IMP divided by labeled ATP + GTP + UTP. The NBI was used as a compact description of the relative labeled IMP and canonical nucleotide pools. It is not a direct measure of enzymatic flux, PRPP availability or nucleotide synthesis rate.

### Statistical analysis

All analyses were performed in Python 3.14 and R 4.3.2. Pools and pathway scores were analyzed on the log₂ scale; fold changes are ratios of geometric means. Proportions and composite indices were analyzed on their original scale. Hedges’ g was calculated for tumor treatment contrasts, and 95% confidence intervals were obtained from the corresponding linear models where estimable [22].

For UK131, we fitted response ∼ tissue × treatment with non-tumor tissue and vehicle as reference levels. For UK2035, we fitted response ∼ pembrolizumab × WGP within block CA2. These models reproduce the analytical structure used in the source study and are reported to allow direct comparison.

Because several treatment cells contained only two or three samples, we also performed exact randomization tests. For the UK131 tumor contrast, all 20 assignments of three pembrolizumab and three vehicle labels were enumerated. For the UK131 tissue × treatment interaction, treatment labels were permuted independently within tumor and non-tumor tissue, yielding 400 assignments. For the UK2035 factorial experiment, all 92,400 assignments compatible with cell sizes 3/3/3/2 were enumerated. The two-sided exact P value was the proportion of assignments with an absolute test statistic at least as large as the observed statistic [24]. These tests do not assume normality but are limited by the fixed design; for a 3-versus-3 tumor contrast, the smallest attainable two-sided exact P value is 0.10.

Benjamini–Hochberg q values were calculated within endpoint families: glucose indices, glutamine indices, glucose pathway scores, glutamine pathway scores and UK2035 nucleotide endpoints [23]. Model-based P values are reported alongside exact P values because the two approaches answer related but not identical questions. Exact tests were performed as a sensitivity analysis after the reanalysis was specified, and all q values are therefore descriptive rather than confirmatory.

The macrophage fingerprint comparison used 20,000 random re-orderings of one effect vector and is reported only in the Supporting Information. No inferential claim was based on this comparison.

### Ethics statement

This study is a secondary analysis of de-identified, publicly deposited data. No new participants were recruited, no new specimens were collected and no identifiable private information was accessed. The ethics approvals and consent procedures for the original tissue collection are reported in the source publication [8].

### Data and code availability

The source dataset is publicly available from Dryad at https://doi.org/10.5061/dryad.n5tb2rbwc [9]. Processed analysis tables and the complete analysis code are provided as Supporting Information. The reanalysis scripts reproduce the quality-control summaries, model-based estimates, exact randomization tests and figures from the deposited tables.

## Results

### Dataset composition and replicate variability

The public deposit contained 54 sample records across five workbooks. After design filtering, 38 tissue samples from UK131 and UK2035 were retained for the primary analysis, and 16 macrophage samples were retained only for descriptive context (Table 1; Figure 1). UK131 contributed matched tumor and non-tumor tissue for both glucose and glutamine tracing. UK2035 contributed 11 samples in the complete CA2 factorial block and three additional vehicle-treated samples in block CA.

**Figure 1.**
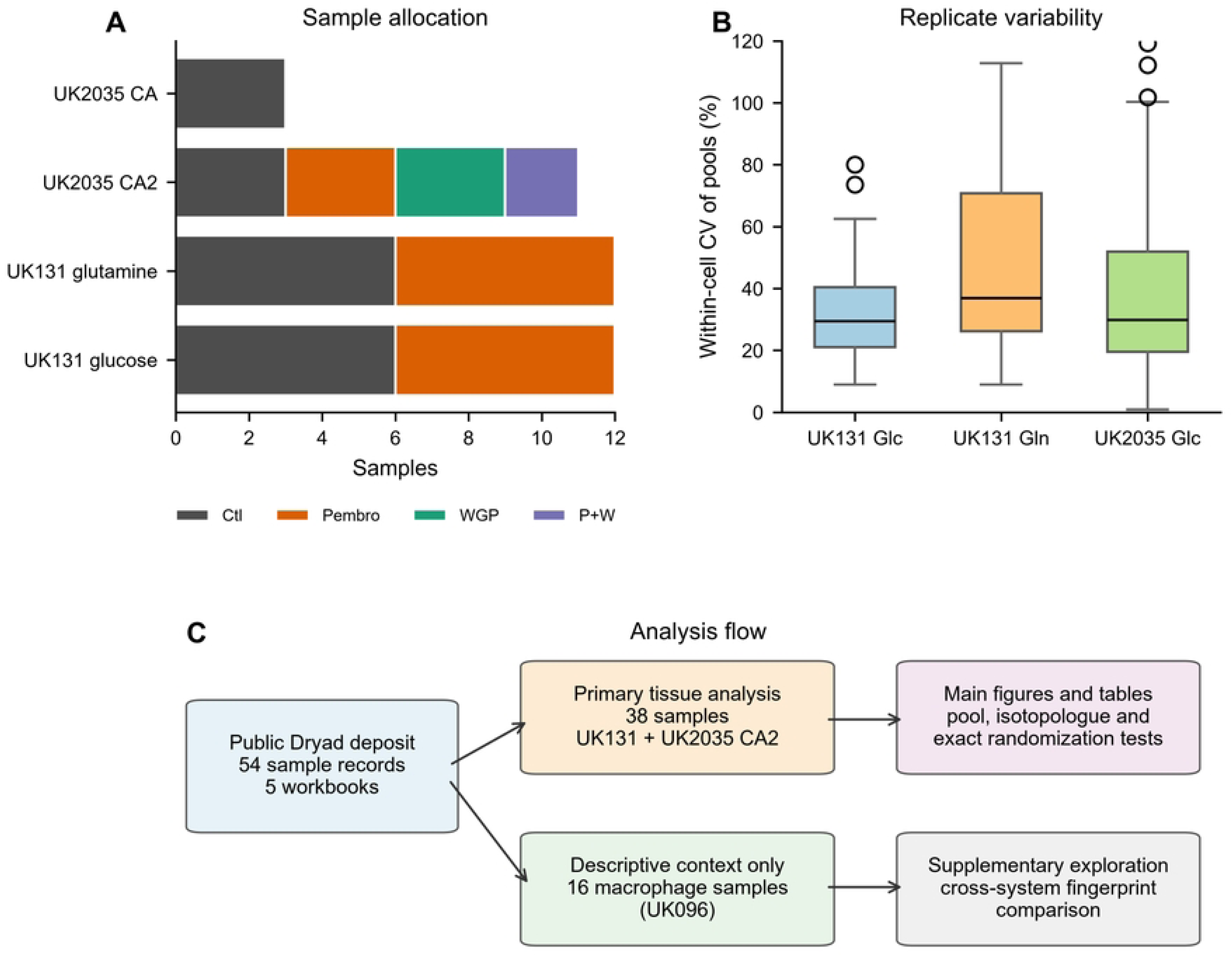
Study design, replicate variability and analysis flow. (A) Sample allocation across the two patients and tracer experiments. (B) Within-cell coefficients of variation for metabolite pools. Boxes show the median and interquartile range; whiskers extend to 1.5 times the interquartile range. (C) Flow from the public deposit to the primary tissue analysis and the descriptive macrophage comparison. U-¹³C₆-glucose and U-¹³C₅,¹⁵N₂-glutamine are abbreviated as glucose and glutamine tracers in panel A. WGP, whole glucan particle β-glucan.

**Table 1.** Experimental design and samples used in the reanalysis.

| Experiment | Patient | Tracer | Tissue and treatment cells | Metabolites analyzed | Samples | Role |
|---|---|---|---|---|---|---|
| UK131 glucose | UK131 | U-<sup>13</sup>C<sub>6</sub>-glucose | CA vehicle 3, CA pembrolizumab 3; NC vehicle 3, NC pembrolizumab 3 | 8 | 12 | Primary |
| UK131 glutamine | UK131 | U-<sup>13</sup>C<sub>5</sub>,<sup>15</sup>N<sub>2</sub>-glutamine | CA vehicle 3, CA pembrolizumab 3; NC vehicle 3, NC pembrolizumab 3 | 10 | 12 | Primary |
| UK2035 CA2 | UK2035 | U-<sup>13</sup>C<sub>6</sub>-glucose | Vehicle 3, pembrolizumab 3, WGP 3, pembrolizumab + WGP 2 | 24 | 11 | Primary factorial |
| UK2035 CA | UK2035 | U-<sup>13</sup>C<sub>6</sub>-glucose | Vehicle 3 | 24 | 3 | Excluded from factorial inference |
| UK096 macrophages | UK096 | Glucose or glutamine | M0, M1, M2, M2 + WGP; 2 per condition | 8 or 10 | 16 | Descriptive only |

Within-cell CVs for metabolite pools were substantial. Median CVs were 29.5% for UK131 glucose, 36.8% for UK131 glutamine and 29.8% for UK2035 glucose, with upper quartiles of 40.4%, 70.8% and 51.9%, respectively (Figure 1B). Coverage differed systematically between experiments because each workbook used a different metabolite panel. These features limit the precision of all treatment estimates and make complete-case, per-endpoint analysis preferable to imputation.

CA, tumor tissue; NC, non-tumor tissue; WGP, whole glucan particle β-glucan. UK2035 block CA2 contained all four treatment groups and was used for factorial inference. Block CA contained vehicle-treated tissue only and was not combined with CA2 because absolute pool scale differed between blocks.

### Pembrolizumab-associated glucose labeling in UK131

In UK131 tumor tissue, labeled non-oxidative PPP intermediates increased from a geometric mean of 0.63 to 1.16 μmol/g protein, a 1.84-fold change (model Welch P = 0.036; model interaction P = 0.145). The exact tumor treatment P value was 0.10, the exact tissue × treatment interaction P value was 0.16, and the Benjamini–Hochberg q value across glucose pathway scores was 0.32. The non-oxidative PPP share increased 1.25-fold (exact interaction P = 0.20; q = 0.36). The oxidative PPP score decreased 0.73-fold, but the estimate was imprecise (model P = 0.42).

The weighted isotopic-scrambling index increased 1.31-fold (0.0509 to 0.0665; model P = 0.0015; model interaction P = 0.012; model-based FDR = 0.0074). In the exact analysis, the tumor treatment P value was 0.10 and the tissue × treatment interaction P value was 0.04, with q = 0.20 across glucose indices. Thus, the scrambling result was the most consistent glucose-related signal in the model-based analysis, but it did not remain below q < 0.05 after exact randomization and multiplicity correction.

Individual non-oxidative PPP metabolites moved in the same direction, but none had a model-based FDR below 0.05. The two composite summaries—non-oxidative PPP share and weighted scrambling—captured the same pattern without adding independent evidence. The data therefore support a directional, tumor-enriched glucose-labeling pattern, not a statistically robust or flux-defined PPP activation (Figure 2).

**Figure 2.**
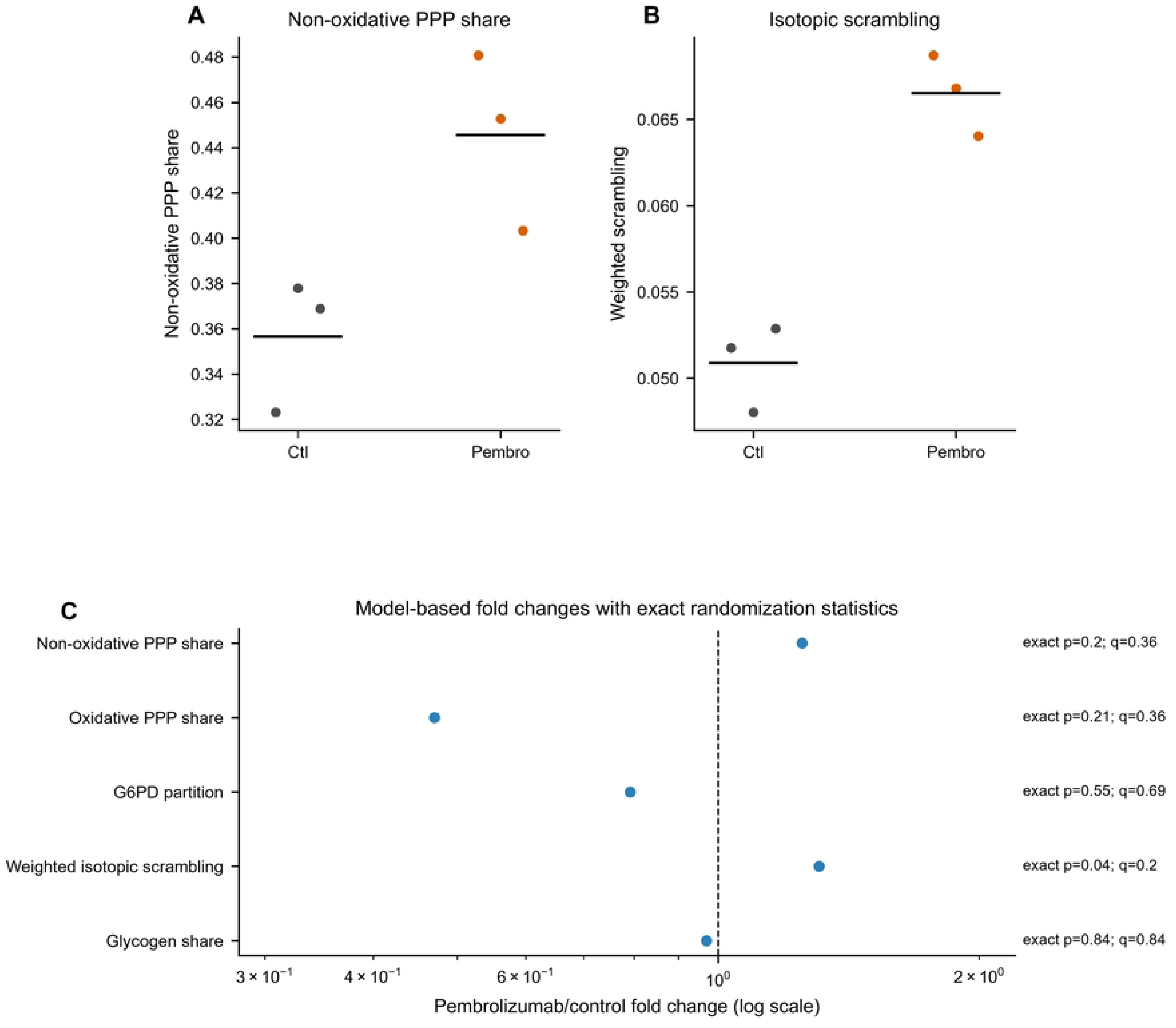
Glucose labeling response to pembrolizumab in UK131 tumor tissue. (A) Non-oxidative PPP share in vehicle- and pembrolizumab-treated tumor tissue. (B) Weighted isotopic-scrambling index. Horizontal lines indicate arithmetic means. (C) Model-based fold changes for glucose pathway indices. Exact P values are two-sided randomization tests; q values are Benjamini–Hochberg-adjusted within the glucose-index family. The x-axis is logarithmic. Pembrolizumab/control ratios were calculated from arithmetic means for display; model-based fold changes and exact statistics are reported in Table 2 and S3 Table.

**Table 2.** Primary labeling estimates and exact randomization statistics.

| **Experiment and endpoint** | **Contrast** | **n** | **Fold change** | **Model P** | **Exact P** | **BH q** | **Interpretation** |
|---|---|---|---|---|---|---|---|
| UK131 non-oxidative PPP score | Pembrolizumab vs vehicle in tumor | 3/3 | 1.84 | 0.036 | 0.10 tumor; 0.16 interaction | 0.32 | Directional only |
| UK131 non-oxidative PPP share | Pembrolizumab vs vehicle in tumor | 3/3 | 1.25 | 0.039 | 0.20 interaction | 0.36 | Directional only |
| UK131 weighted isotopic scrambling | Pembrolizumab vs vehicle in tumor | 3/3 | 1.31 | 0.0015 | 0.04 interaction | 0.20 | Hypothesis-generating |
| UK131 GSH diversion | Pembrolizumab vs vehicle in tumor | 3/3 | 1.66 | 0.019 | 0.09 interaction | 0.22 | Hypothesis-generating |
| UK2035 NBI | Pembrolizumab + WGP interaction | 3/3/3/2 | 3.64 | 3.2 × 10<sup>−4</sup> | 0.016 | 0.097 | Hypothesis-generating |
| UK2035 labeled IMP | Pembrolizumab + WGP interaction | 3/3/3/2 | 1.84 | 0.072 | 0.049 | 0.147 | Hypothesis-generating |
| UK2035 labeled ATP | Pembrolizumab + WGP interaction | 3/3/3/2 | 0.53 | 0.95 | 0.95 | 0.95 | Imprecise |
| UK2035 labeled GTP | Pembrolizumab + WGP interaction | 3/3/3/2 | 0.31 | 0.86 | 0.87 | 0.95 | Imprecise |
| UK2035 labeled UTP | Pembrolizumab + WGP interaction | 3/3/3/2 | 0.30 | 0.75 | 0.77 | 0.95 | Imprecise |
| UK2035 labeled PRPP | Pembrolizumab + WGP interaction | 3/3/3/2 | 0.28 | 0.80 | 0.82 | 0.95 | Imprecise |

### Pembrolizumab-associated glutamine labeling in UK131

The GSH-diversion index increased from 0.158 to 0.262 in tumor tissue, a 1.66-fold change (model Welch P = 0.019; model interaction P = 0.025; model-based FDR = 0.076). The exact tumor treatment P value was 0.10, the exact interaction P value was 0.09, and q was 0.22 across glutamine indices. Non-tumor tissue showed little change (1.09-fold). The total GSH pool did not increase convincingly: the geometric-mean model estimate was 1.35-fold (P = 0.62), and sample-level values were highly variable.

Other glutamine fate estimates were compatible with unchanged or lower TCA-cycle entry, but all intervals were wide. The TCA labeling yield changed 1.36-fold (exact interaction P = 0.21), aspartate yield 0.63-fold (exact interaction P = 0.11) and malic-enzyme yield 0.50-fold (exact interaction P = 0.39). None of the pathway-level glutamine scores had an exact q value below 0.05.

The glutamine data therefore provide a second directional pattern—greater partitioning of labeled glutamine toward GSH without clear pool expansion—but do not establish increased GSH turnover, redox stress or a specific enzymatic mechanism. The zero value in one aspartate-yield sample also illustrates why ratio-based indices require cautious interpretation in very small groups (Figure 3).

**Figure 3.**
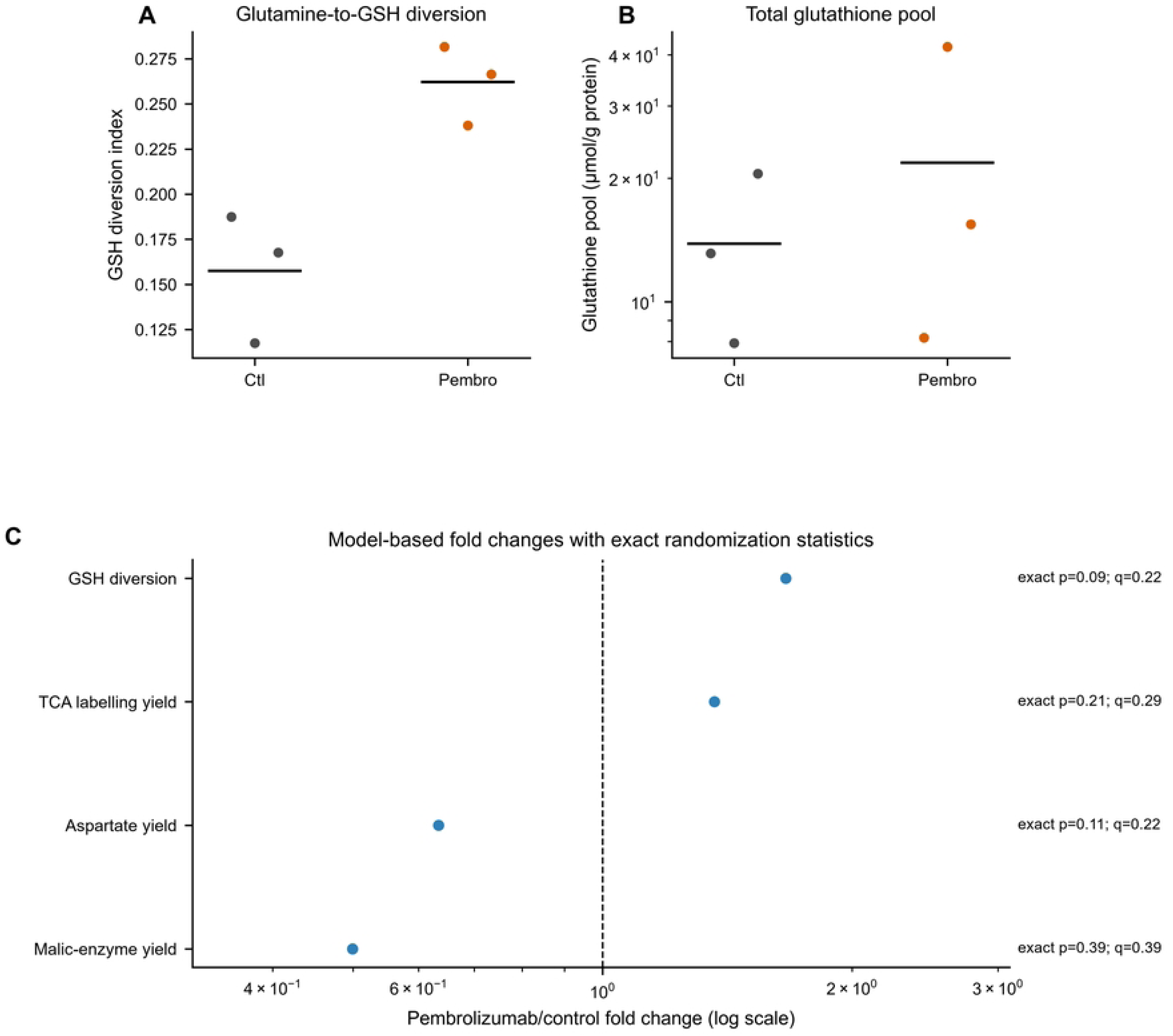
Glutamine labeling and glutathione diversion in UK131 tumor tissue. (A) GSH-diversion index in vehicle- and pembrolizumab-treated tumor tissue. (B) Total glutathione pool on a logarithmic y-axis. Horizontal lines indicate arithmetic means. (C) Model-based fold changes for glutamine fate indices. Exact P values are two-sided randomization tests; q values are Benjamini–Hochberg-adjusted within the glutamine-index family. The x-axis is logarithmic.

### Pembrolizumab × WGP factorial experiment in UK2035

Within block CA2, the NBI was 0.00943 in vehicle, 0.01038 with pembrolizumab, 0.00851 with WGP and 0.03436 with the combination. The combination/control ratio was 3.64. The parametric factorial interaction was β = 0.0249 (P = 3.2 × 10⁻⁴; model-based FDR = 0.0038), whereas the exact randomization interaction P value was 0.016 and the q value across nucleotide endpoints was 0.097.

Labeled IMP increased 1.84-fold with the combination (model interaction P = 0.072; exact P = 0.049; q = 0.147). Labeled ATP, GTP, UTP and PRPP were lower in the combination arm (0.53-, 0.31-, 0.30- and 0.28-fold, respectively), but their exact interaction P values ranged from 0.77 to 0.95. Glycolysis and lactate were not lower in the combination arm, but the estimates were too imprecise to support a selective block in nucleotide synthesis.

These results reproduce the direction of the source study’s combination-associated nucleotide pattern and show that a compact NBI can summarize it. They do not establish a nucleotide biosynthesis bottleneck: the NBI can change because of altered precursor supply, reduced interconversion, nucleotide degradation, dilution by unlabeled material, cell death or analytical recovery. The exact analysis reduced the strength of the evidence substantially relative to the parametric model (Figure 4; Table 2).

**Figure 4.**
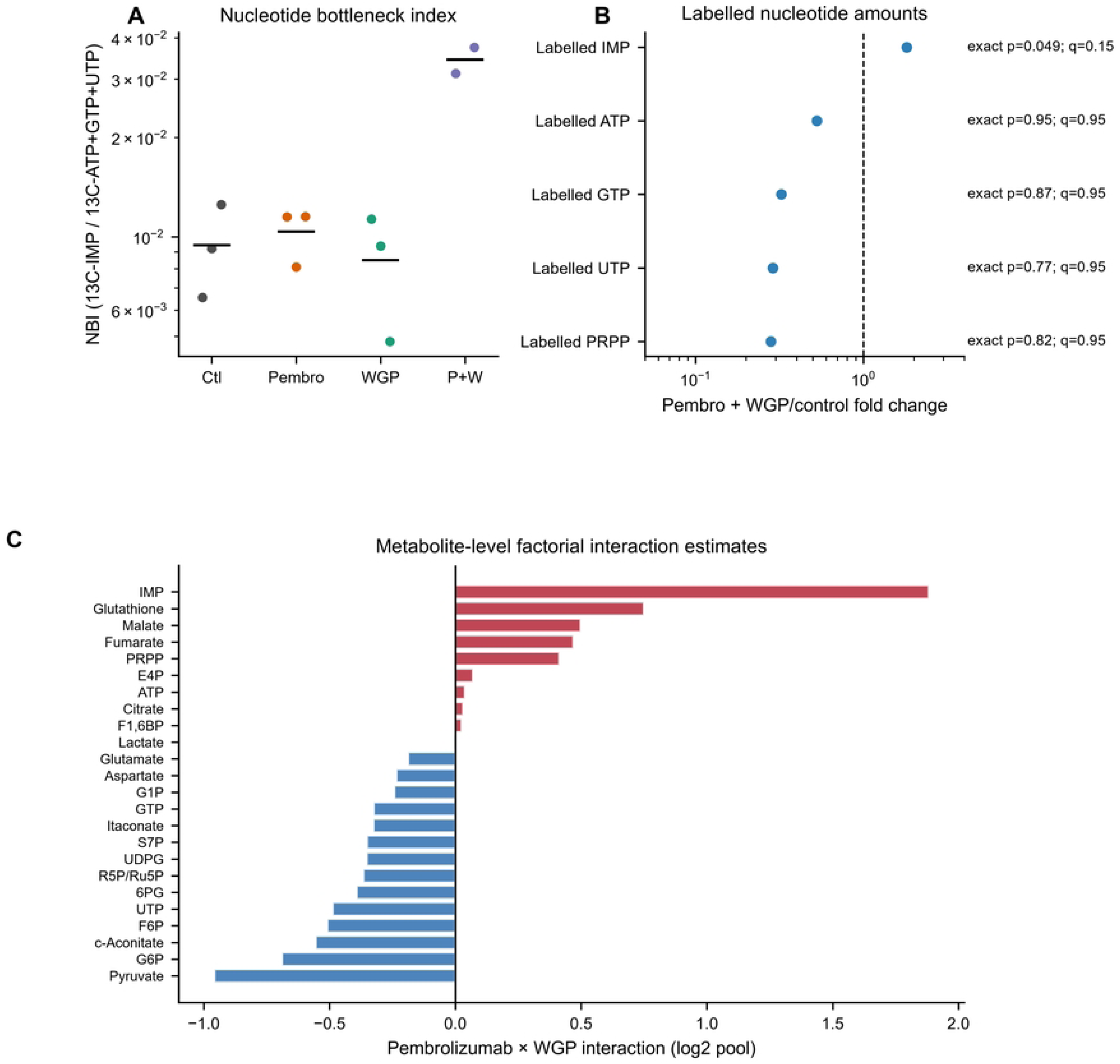
Pembrolizumab × WGP factorial experiment in UK2035 tumor tissue. (A) Nucleotide bottleneck index by treatment arm on a logarithmic y-axis. Horizontal lines indicate arithmetic means. (B) Labeled IMP, ATP, GTP, UTP and PRPP in the combination arm relative to vehicle. Exact P values are two-sided randomization tests for the factorial interaction; q values are Benjamini–Hochberg-adjusted across nucleotide endpoints. (C) Metabolite-level pembrolizumab × WGP interaction estimates from log₂-scale linear models. Positive values indicate a greater combination effect than expected from the single-agent effects. WGP, whole glucan particle β-glucan.

Model P values are from log₂-scale linear models analogous to the source analysis. Exact P values are two-sided randomization tests that enumerate all assignments compatible with the fixed design. For UK131, the smallest attainable two-sided exact P value for a 3-versus-3 tumor contrast is 0.10. BH q values are calculated within endpoint families and are descriptive. No endpoint reached q < 0.05 in the exact analysis. NBI, nucleotide bottleneck index; PPP, pentose phosphate pathway; GSH, glutathione; WGP, whole glucan particle β-glucan.

### Comparison of model-based and exact inference

Across the principal endpoints, exact randomization P values were consistently larger than model-based interaction P values (Figure 5). The difference was largest for the NBI (3.2 × 10⁻⁴ versus 0.016), weighted scrambling (0.012 versus 0.040) and GSH diversion (0.025 versus 0.090). This pattern is expected when normal-theory tests are applied to very small treatment cells, but its magnitude is large enough to change the interpretation of the study.

**Figure 5.**
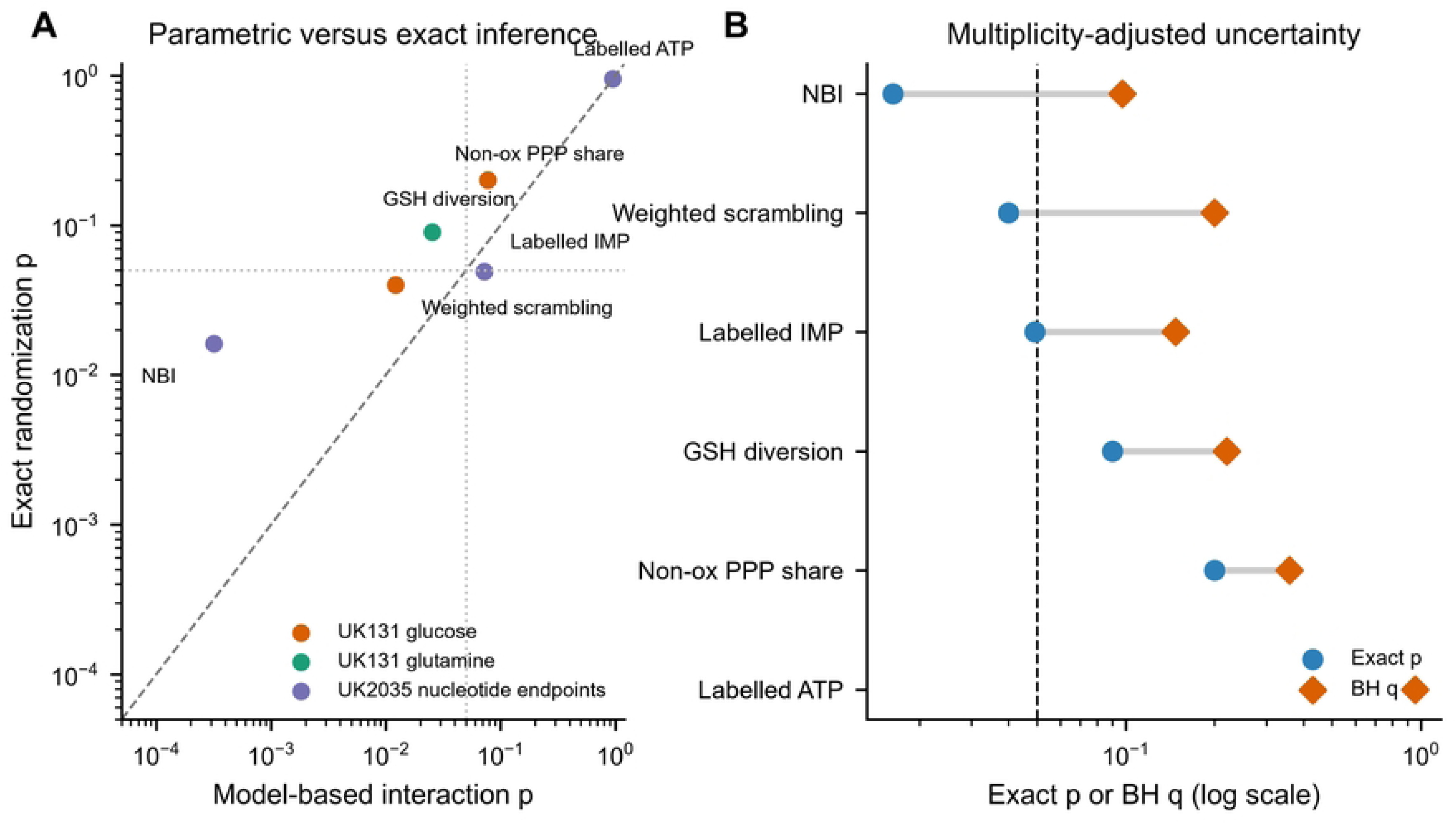
Comparison of model-based and exact randomization inference. (A) Model-based interaction P values against exact randomization P values for six principal endpoints. The dashed line indicates equality; points below the line indicate larger exact P values. Dotted lines mark P = 0.05. (B) Exact P values and Benjamini–Hochberg q values for the same endpoints. q values are family-specific and are shown together only for visual comparison. No endpoint reached q < 0.05.

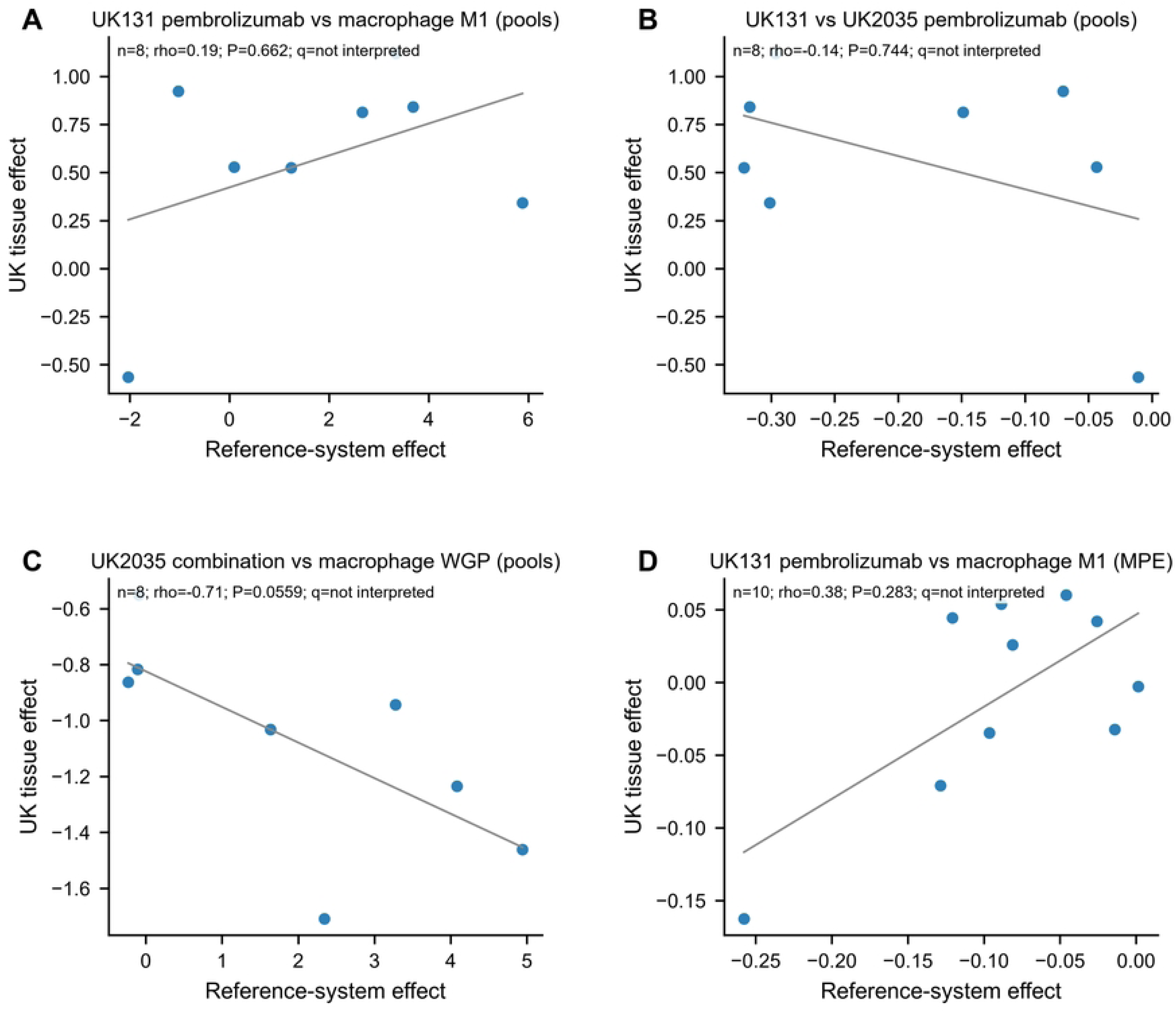

The exact tests also make the design limitation explicit: a two-sided 3-versus-3 comparison has only 20 possible assignments, so no tumor treatment P value can be smaller than 0.10 regardless of effect size. The factorial UK2035 design permits smaller P values, but only two samples were available in the combination arm, and correction across nucleotide endpoints moved the NBI q value to 0.097. These constraints cannot be resolved by reanalysis; they require additional biological replicates or orthogonal experiments.

### Exploratory cross-system comparison

The 14 macrophage–tissue comparisons are reported in Figure S1 and S4 Table. The only nominally significant Spearman correlation was an inverse relationship between the UK2035 combination effect and the macrophage WGP response across eight glucose metabolites (ρ = −0.79; permutation P = 0.026), but its Benjamini–Hochberg q value across the 14 correlations was 0.37. The direction and magnitude of this correlation were sensitive to the small metabolite set and were not interpreted as evidence of opposing biological programs. No cross-system comparison was used to support a primary conclusion.

## Discussion

This independent reanalysis reproduced the direction of the source study’s three principal metabolic patterns: tumor-enriched non-oxidative PPP labeling and isotopic scrambling in UK131, glutamine-derived GSH diversion in UK131, and a combination-associated nucleotide pattern in UK2035. It also showed that the statistical support for these patterns is considerably weaker than conventional model-based P values suggest. The most defensible conclusion is therefore not that a nucleotide bottleneck or a specific PPP mechanism has been demonstrated, but that public SIRM data can be recomputed transparently and that the resulting treatment effects require larger, orthogonal studies.

The glucose results are consistent with increased reversible sugar-phosphate exchange and non-oxidative PPP labeling in the tumor compartment. However, the data do not distinguish transketolase/transaldolase cycling from altered labeling transit time, changes in cell composition, tissue viability or precursor dilution. The oxidative PPP score and G6PD partition did not increase, but their confidence intervals were wide. A change confined to non-oxidative intermediates is compatible with ribose-5-phosphate supply or glycogen-linked cycling, as discussed in the source study, but the present dataset cannot identify the responsible enzyme or flux [14,15].

The glutamine results show a similar boundary. A higher fraction of labeled glutamine recovered in GSH, without a clear increase in total GSH, is compatible with increased GSH turnover, but it is also compatible with altered precursor labeling, compartmentation or sample recovery. The source study’s immune and tissue-damage measurements are essential for interpreting this pattern and were not part of the present data-level reanalysis. The reanalysis therefore refines the quantitative description of the published observation without adding a new mechanistic layer.

The UK2035 NBI is best regarded as an analysis-derived summary of labeled IMP relative to canonical nucleotides. The combination arm showed a large point estimate, but the exact interaction P value was 0.016 and the family-adjusted q value was 0.097. Labeled ATP, GTP, UTP and PRPP were lower with wide uncertainty, and labeled IMP was higher with an exact P value of 0.049. These estimates are compatible with a combination-associated nucleotide constraint, but they do not establish a block at PRPP supply, IMP interconversion or downstream nucleotide synthesis. A true flux estimate would require targeted confirmation with stable-isotope flux analysis, nucleotide pool turnover measurements, enzyme activity or genetic perturbation, and orthogonal viability and cell-composition measurements.

The methodological finding is important for the interpretation of small patient-derived tracer studies. With three samples per group, the exact two-sided randomization test for a tumor treatment effect cannot produce P < 0.10, even for a large effect. Parametric tests can produce much smaller P values by invoking distributional assumptions that cannot be checked in such small samples. The discrepancy observed here—particularly for the NBI and GSH-diversion indices—shows why effect sizes, sample-level displays, exact tests and multiplicity correction should be reported together [22–25]. The solution is not to choose the test that produces the smallest P value, but to design experiments with enough biological replication and to prespecify the estimands and composite endpoints.

The cross-system comparison illustrates a related problem. Correlation between treatment-effect vectors based on eight metabolites, two to three replicates per condition and 14 comparisons is unstable. The nominal inverse correlation between the UK2035 combination and macrophage WGP response did not survive multiplicity correction and should not be interpreted as evidence that tumor tissue and macrophages respond in opposite directions. We therefore retain this analysis only as an exploratory supplement.

This study has several limitations. Only two tumor patients were available, and the two patients differed in tissue source (primary lung tumor versus brain metastasis), treatment design and tracer panel. The treatment cells contained two or three samples, replicate CVs were high and coverage was panel-specific. The composite indices were constructed during reanalysis and were not validated in an independent cohort. The exact tests are design-limited, not a substitute for replication. No cytokine, viability, immune-cell composition, enzyme activity or flux measurements were available. Finally, the reanalysis cannot determine whether the observed labeling patterns reflect tumor cells, immune cells or other components of the tissue slice.

More broadly, tumor and immune cells compete for nutrients, and exhaustion-associated mitochondrial stress may further amplify the context dependence of metabolic readouts [26,27,28,29]. These limitations do not make the dataset uninformative. They define the appropriate claim: the public deposit supports directional, patient- and tissue-context-specific metabolic hypotheses that are suitable for prioritization in larger studies. The reanalysis also provides a reproducible template for reusing SIRM data, including exact randomization tests, family-wise q values, sample-level displays and explicit separation of labeling from flux.

## Conclusions

Independent reanalysis of public SIRM data recovered the direction of the source study’s glucose, glutamine and nucleotide-labeling patterns, but exact randomization tests showed that the statistical evidence is limited by the original design. The findings should be used to generate hypotheses about tumor-context metabolic adaptation and combination immunotherapy, not to claim a validated PPP mechanism or nucleotide bottleneck. Larger patient-derived experiments with independent replication and orthogonal functional measurements are required before these metabolic readouts can be considered biomarkers or therapeutic targets.

## Acknowledgments

The authors thank the investigators who generated and deposited the source dataset.

## Author contributions

Zhijie Shang: Conceptualization, Data curation, Formal analysis, Investigation, Writing – original draft, Visualization

Hong Geng: Conceptualization, Supervision, Methodology, Writing – review & editing, Project administration.

## Ethics approval and consent to participate

Not applicable. This study is a secondary analysis of de-identified, publicly deposited data. The original study obtained the ethics approvals and consent reported in the source publication [8].

## Consent for publication

Not applicable.

## Data availability

The source dataset is available from Dryad at https://doi.org/10.5061/dryad.n5tb2rbwc. Processed data and analysis code are provided as Supporting Information.

## Use of artificial intelligence

The authors used AI-assisted tools for language editing and manuscript formatting. The authors reviewed and edited all AI-assisted content and take full responsibility for the manuscript.

## Supporting information

**S1 Fig. Exploratory cross-system comparisons.** Effect vectors for 14 tissue–macrophage comparisons. Each point is a metabolite shared by the two experiments. The displayed Spearman correlations are descriptive; the corresponding Benjamini–Hochberg q value across the 14 correlations was >0.05 for all comparisons. These analyses were not used to support a primary conclusion.

**S1 Table. Full UK131 metabolite and pathway results.** Model-based estimates, confidence intervals, exact randomization P values and family-specific q values for UK131 glucose and glutamine endpoints.

**S2 Table. Full UK2035 metabolite and pathway results.** Model-based factorial estimates, exact randomization P values and family-specific q values for UK2035 block CA2.

**S3 Table. Exact randomization tests.** Enumeration results for principal UK131 and UK2035 endpoints.

**S4 Table. Exploratory cross-system comparisons.** Spearman correlations and permutation P values for the 14 tissue–macrophage comparisons.

**S1 Code and processed data.** Analysis scripts, processed sample-level data and statistics tables required to reproduce the figures and tables.

## References

1. Pardoll DM. The blockade of immune checkpoints in cancer immunotherapy. Nat Rev Cancer. 2012;12(4):252–264. doi:10.1038/nrc3239

2. O’Neill LA, Kishton RJ, Rathmell J. A guide to immunometabolism for immunologists. Nat Rev Immunol. 2016;16(9):553–565. doi:10.1038/nri.2016.70

3. Leone RD, Powell JD. Fueling the revolution: targeting metabolism to enhance immunotherapy. Cancer Immunol Res. 2021;9(3):255–260. doi:10.1158/2326-6066.CIR-20-0791

4. Vander Heiden MG, Cantley LC, Thompson CB. Understanding the Warburg effect: the metabolic requirements of cell proliferation. Science. 2009;324(5930):1029–1033. doi:10.1126/science.1160809

5. DeBerardinis RJ, Lum JJ, Hatzivassiliou G, Thompson CB. The biology of cancer: metabolic reprogramming fuels cell growth and proliferation. Cell Metab. 2008;7(1):11–20. doi:10.1016/j.cmet.2007.10.002

6. DeBerardinis RJ, Chandel NS. Fundamentals of cancer metabolism. Sci Adv. 2016;2(5):e1600200. doi:10.1126/sciadv.1600200

7. Chang CH, Qiu J, O’Sullivan D, Buck MD, Noguchi T, Curtis JD, et al. Metabolic competition in the tumor microenvironment is a driver of cancer progression. Cell. 2015;162(6):1229– 1241. doi:10.1016/j.cell.2015.08.016

8. Fan TW, Higashi RM, Song H, Daneshmandi S, Mahan AL, Purdom MS, et al. Innate immune activation by checkpoint inhibition in human patient-derived lung cancer tissues. eLife. 2021;10:e69578. doi:10.7554/eLife.69578

9. Lane AN, Fan TW, Higashi RM, Song H, Daneshmandi S, Mahan AL, et al. Innate immune activation by checkpoint inhibition in patient-derived lung cancer tissues [dataset]. Dryad. 2021. doi:10.5061/dryad.n5tb2rbwc

10. Bengsch B, Johnson AL, Kurachi M, Odorizzi PM, Pauken KE, Attanasio J, et al. Bioenergetic insufficiencies due to metabolic alterations regulated by the inhibitory receptor PD-1 are an early driver of CD8+ T cell exhaustion. Immunity. 2016;45(2):358–373. doi:10.1016/j.immuni.2016.07.008

11. Lane AN, Fan TW. Regulation of mammalian nucleotide metabolism and biosynthesis. Nucleic Acids Res. 2015;43(4):2466–2485. doi:10.1093/nar/gkv047

12. Stern A, Fokra M, Sarvin B, Alrahem AA, Lee WD, Aizenshtein E, et al. Inferring mitochondrial and cytosolic metabolism by coupling isotope tracing and deconvolution. Nat Commun. 2023;14:7525. doi:10.1038/s41467-023-42824-z

13. Antoniewicz MR. A guide to 13C metabolic flux analysis for the cancer biologist. Exp Mol Med. 2018;50(4):1–13. doi:10.1038/s12276-018-0060-y

14. Patra KC, Hay N. The pentose phosphate pathway and cancer. Trends Biochem Sci. 2014;39(8):347–354. doi:10.1016/j.tibs.2014.06.005

15. Stincone A, Prigione A, Cramer T, Wamelink MM, Campbell K, Cheung E, et al. The return of metabolism: biochemistry and physiology of the pentose phosphate pathway. Biol Rev Camb Philos Soc. 2015;90(3):927–963. doi:10.1111/brv.12140

16. Schafer FQ, Buettner GR. Redox environment of the cell as viewed through the redox state of the glutathione disulfide/glutathione couple. Free Radic Biol Med. 2001;30(11):1191– 1212. doi:10.1016/S0891-5849(01)00480-4

17. Yang L, Venneti S, Nagrath D. Glutaminolysis: a hallmark of cancer metabolism. Annu Rev Biomed Eng. 2017;19:163–194. doi:10.1146/annurev-bioeng-071516-044546

18. O’Neill LA, Artyomov MN. Itaconate: the poster child of metabolic reprogramming in macrophage function. Nat Rev Immunol. 2019;19(5):273–281. doi:10.1038/s41577-019-0128-5

19. Ledwith AE, Prendeville H, Horneck Johnston CJ, Cunningham SP, McGrath JP, Corkish C, et al. Yeast β-glucan supplementation supports immunometabolic anti-tumor responses and reverses obesity-induced dysfunction via trained hematopoiesis. Cell Rep. 2026;45:117648. doi:10.1016/j.celrep.2026.117648

20. Netea MG, Joosten LA, Latz E, Mills KH, Natoli G, Stunnenberg HG, et al. Trained immunity: a program of innate immune memory in health and disease. Science. 2016;352(6284):aaf1098. doi:10.1126/science.aaf1098

21. O’Neill LA, Kishton RJ, Rathmell J. Immunometabolism governs dendritic cell and macrophage function. J Exp Med. 2016;213(1):15–23. doi:10.1084/jem.20151570

22. Lakens D. Calculating and reporting effect sizes to facilitate cumulative science: a practical primer for t-tests and ANOVAs. Front Psychol. 2013;4:863. doi:10.3389/fpsyg.2013.00863

23. Benjamini Y, Hochberg Y. Controlling the false discovery rate: a practical and powerful approach to multiple testing. J R Stat Soc Series B Stat Methodol. 1995;57(1):289–300. doi:10.1111/j.2517-6161.1995.tb02031.x

24. Phipson B, Smyth GK. Permutation P-values should never be zero: calculating exact P-values when permutations are randomly drawn. Stat Appl Genet Mol Biol. 2010;9:Article39. doi:10.2202/1544-6115.1585

25. Button KS, Ioannidis JP, Mokrysz C, Nosek BA, Flint J, Robinson ES, et al. Power failure: why small sample size undermines the reliability of neuroscience. Nat Rev Neurosci. 2013;14(5):365–376. doi:10.1038/nrn3475

26. Mills EL, Kelly B, Logan A, Costa AS, Varma M, Bryant CE, et al. Succinate dehydrogenase supports metabolic repurposing of mitochondria to drive inflammatory macrophages. Cell. 2016;167(2):457–470.e13. doi:10.1016/j.cell.2016.08.064

27. Binnewies M, Roberts EW, Kersten K, Chan V, Fearon DF, Merad M, et al. Understanding the tumor immune microenvironment (TIME) for effective therapy. Nat Med. 2018;24(5):541– 550. doi:10.1038/s41591-018-0014-x

28. Wherry EJ, Kurachi M. Molecular and cellular insights into T cell exhaustion. Nat Rev Immunol. 2015;15(8):486–499. doi:10.1038/nri3862

29. Scharping NE, Rivadeneira DB, Menk AV, Vignali PDA, Ford BR, Rittenhouse NL, et al. Mitochondrial stress induced by continuous stimulation under hypoxia rapidly drives T cell exhaustion. Nat Immunol. 2021;22(2):205–215. doi:10.1038/s41590-020-00834-9

